# How accurate are our near reading charts? An assessment of 19 charts against ISO standards

**DOI:** 10.64898/2026.01.29.26345152

**Authors:** Timothy I. Murphy, Jingyi Chen, Myra Leung

## Abstract

**Purpose:** Measuring near vision provides clinicians with valuable insight into visual function. There is limited information on the accuracy of available reading charts frequently used in community practice. This study aimed to measure internationally available reading charts to determine how they compare to international standards.

**Methods:** Commercially and device manufacturer-provided reading charts available in community optometry clinics were scanned at 600 dots per inch. Gaussian adaptive threshold was used to facilitate repeatable measurements. X- heights of letters were measured independently by three researchers. Contrast levels and line spacing were also estimated. Results for each chart were compared with ISO Standards. Intraclass correlation coefficient was used to assess intergrader agreement.

**Results:** Of the 19 reading charts that were measured, only one chart (5.26%) had text sizes that were all within tolerance. There was high variability in size observed between charts. Twelve charts (63.2%) used serif fonts and seven used sans-serif (36.8%). Text on serif charts tended to be smaller than required (µ=−9.63%) compared to sans-serif (µ=+4.96%). All charts met the line spacing requirements and minimum required contrast level; however, some charts were printed on laminated or satin plastic which does not meet the standard of using a matte surface. There was high interrater agreement (ICC(2,1) = 1.00), indicating a highly repeatable measurement technique.

**Conclusion:** This study found that the tested reading charts displayed significant variability in text size. Although some charts had more lines of text within size tolerances than others, none met all the requirements of the International Standard. Clinicians and researchers should take care when interpreting changes in near reading acuity when multiple charts have been used, especially as part of shared care models or when monitoring progressive vision changes. A free UC/UWA Reading Chart has been developed as a result of this study, which conforms to the ISO Standard.

**Key points:**

1. None of the measured reading charts met the requirements of the ISO 7921:2024 standard.
2. There is high variability in text size between reading charts.
3. A new chart, the UC/UWA Reading Chart, has been developed to conform to the ISO standards.

## Introduction

Near reading acuity charts are integral to functional vision assessment and are commonplace in community ophthalmic practice. While there are national and international guidelines for reading chart development [1, 2], standardisation research has largely focused on reading speed, word difficulty and correlation with distance acuity on specially designed and validated products [3–5]. In community practice, however, the use of non-specialised charts, including those given to practices by lens manufacturers, is common. There is limited evidence regarding the accuracy of these charts, especially with respect to text size which can be affected by font type [6]. Chart differences can be problematic where a clinician uses multiple charts, such as simple text for children, more complex text for adults or larger text for low vision, or where different charts are used in different rooms in the same clinic. Standardisation and interoperability of near reading charts are necessary to effectively bridge the gap between clinical research outcomes and real-world practice [6, 7]. This allows for accurate monitoring of functional vision for disease progression, which is especially important for low vision management and the prescription of magnification aids [8].

International standard ISO 7921:2024 [1], first published in 2024, defines parameters and restrictions for font, line spacing, type of text, luminance and contrast, and text size determination and progression. Text size is referenced to x-height, defined as the height of a lowercase ‘x’ without ascenders and descenders (Fig. 1) in millimetres. Text size is defined using Reading Acuity Determination (RAD) at a set distance, which is equivalent to the Minimum Angle of Resolution (MAR) at that distance where the letter size is the height of a lowercase ‘x’, presented as the logarithm of RAD (logRAD). However, near reading charts used in community optometry are often labelled using logMAR, even though logMAR is based on the width of an individual line and mainly used for distance letter acuity charts. Both units can be converted to Snellen equivalents, with the Snellen equivalent of logMAR equal to the Snellen equivalent of logRAD. Hence, for the purposes of this study, the two units have been directly compared despite having different definitions. Deviations of ± 5% for sizes greater than −0.20 logRAD are permitted, or ±10% for smaller sizes. Reading acuity grades can be recorded as logRAD (logMAR), Decimal, M size, N size and/or reduced Snellen fractions. The details of these units are outside of the scope of this paper, though work by Radner [4, 6] provides in-depth analyses for interested readers.

**Fig. 1:**
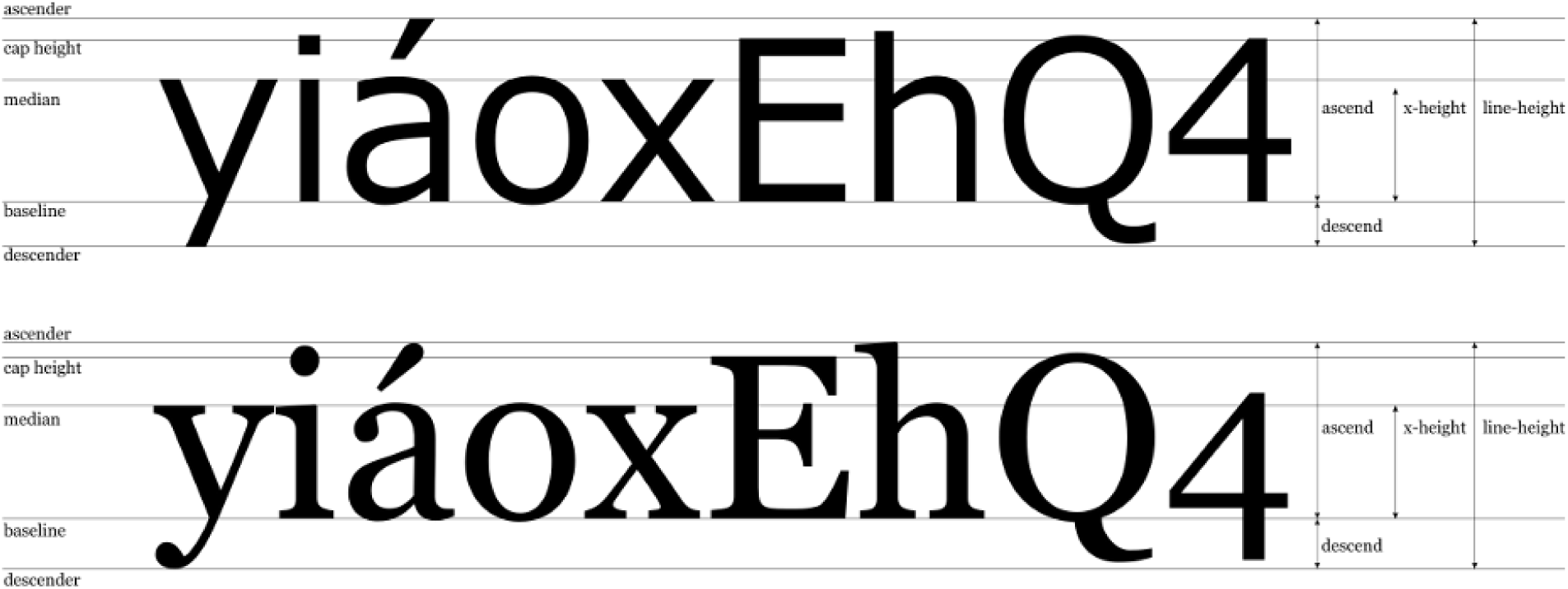
Font measurement standards for sans-serif (top) and serif (bottom) fonts. Image by DekorMeló, Public domain, via Wikimedia Commons.

The standard requires charts to include text at size 0.00 logRAD (or equivalent) with smaller and larger text permitted provided all increments of 0.10 log units are included. Text must have a minimum contrast of 85%, with line spacing no greater than 1.5.

This study aims to assess whether various near reading acuity charts used in clinical practice meet the size requirements and line spacing stipulated in the International standard ISO 7921:2024. First, the accuracy and consistency of text size labels are examined when compared to the standard. Next, text sizes are measured to calculate deviation from the standard, with serif and sans-serif fonts analysed separately. Line spacing is then estimated based on these text size measurements before text contrast is assessed.

## Methods

Internationally manufactured near reading acuity charts, available commercially or provided by ophthalmic device manufacturers, were scanned, measured, and compared with the ISO 7921:2024 standard [1]. These charts were selected through convenience sampling, available to the researchers at universities and their affiliated optometry clinics in Australia, New Zealand and Hong Kong. Charts were scanned at 600 dots per inch (dpi) using a Konica Minolta C450i multi-function printer (Konica Minolta Inc., Tokyo, Japan) and saved as JPEG images. This provided an image with each pixel representing 0.0423mm, allowing letters to be measured in pixels and converted to millimetres.

Letters without curves were measured for each text size on each chart. As many charts did not contain the letter ‘x’ in all sizes, lowercase letters without curves of the same height were measured (‘x’, ‘v’, ‘w’ and ‘z’). Capital letters without curves were also measured (‘A’, ‘E’, ‘F’, ‘H’, ‘I’, ‘K’, ‘L’, ‘M’, ‘N’, ‘T’, ‘V’, ‘W’, ‘X’, ‘Y’, ‘Z’), allowing the cap height to x-height ratio to be calculated and used to derive the x-height where no appropriate lowercase letters were present.

To facilitate objective and repeatable measurements, adaptive thresholding using Gaussian means [9] was used to convert images to binary black and white (Fig. 2). This removed grey pixels, minimising uncertainty when determining the position of letter edges.

**Fig. 2:**
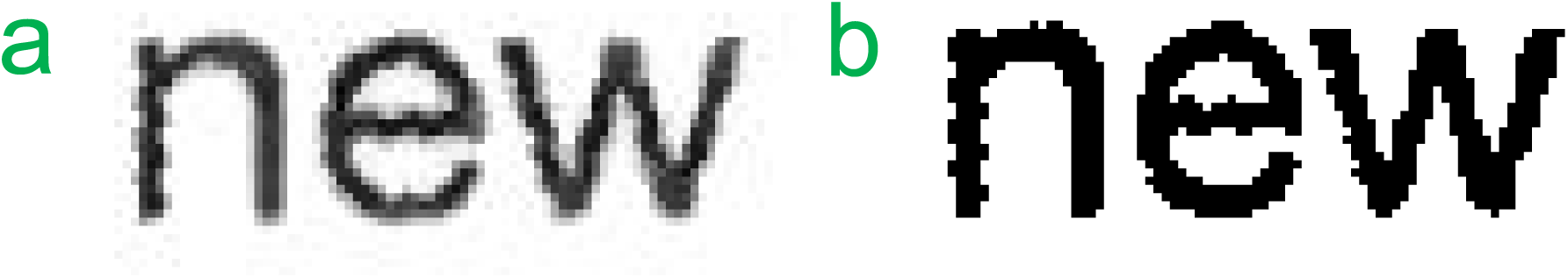
Example of adaptive thresholding applied to 5pt font scanned at 600 dpi. a: original scan. b: scan with adaptive thresholding applied.

The accuracy of this technique was verified by scanning a 20-centimetre (cm) ruler, whose length was verified using callipers, and applying adaptive thresholding to this scan. When oriented vertically, this process resulted in a ruler length of 4724.4 pixels, which at 0.0423 mm per pixel is 20.00cm. This measurement was 4730.0 pixels horizontally, resulting in a measurement error of 0.12% in the horizontal axis. All charts were scanned vertically. More information about this verification procedure is available in the supplementary material.

Letter height was calculated by finding the top and bottom y-coordinate of the letter from the threshold-processed image. From this, the pixel height of the letter was found and converted into millimetres. Each of the three authors selected and measured three uppercase and three lowercase non-curved letters for each font size, or less where three were not available, resulting in up to nine x-height and nine cap height measurements for each size on each chart. No blinding mechanism was undertaken. Graders were free to choose letters they felt had the clearest upper and lower boundaries within the character limitations noted earlier. Mean x-height measurements for each text size were analysed, with the deviation from the expected size calculated as a percentage and compared to the deviation allowed in the standard. Two-way random, single score Intraclass correlation coefficient (ICC(2,1)) was used to test inter-grader agreement [10] using the Pingouin statistics package in Python [11]. The ISO standard conversion table, identical to that produced by Radner et al. [6], was used to compare measurements against the expected values for each unit (Table 1). Serif and sans-serif charts were analysed separately. Where multiple but inconsistent units were provided for the same text (e.g. N8 and −0.10 logMAR), the discrepancy was noted and the unit closest to the measured x-height was used. Total measurement error was defined as the measurement error of the pixel counting method (1 pixel) plus the mean standard deviation of measurements from each grader for each chart text size. Standard deviation data are available in the supplementary material.

**Table 1:** Near acuity notations based upon the x-height, adapted from Radner et al. [6].

| Reading acuity angle<br>min of arc | x-height<br>mm | Near reading acuity grades at 40cm |  |  |  |  |  |
| --- | --- | --- | --- | --- | --- | --- | --- |
|  |  | logRAD | Decimal reading acuity | M size | N size | Reduced Snellen fraction |  |
| 20.0 | 11.61 | +1.30 | 0.05 | 8.00 | 60 | 20/400 | 6/120 |
| 15.8 | 9.22 | +1.20 | 0.063 | 6.30 | 48 | 20/320 | 6/95 |
| 12.6 | 7.33 | +1.10 | 0.08 | 5.00 | 36 | 20/250 | 6/75 |
| 10.0 | 5.82 | +1.00 | 0.10 | 4.00 | 30 | 20/200 | 6/60 |
| 7.94 | 4.62 | +0.90 | 0.125 | 3.20 | 24 | 20/160 | 6/48 |
| 6.31 | 3.67 | +0.80 | 0.16 | 2.50 | 18 | 20/126 | 6/38 |
| 5.01 | 2.92 | +0.70 | 0.20 | 2.00 | 14 | 20/100 | 6/30 |
| 3.98 | 2.32 | +0.60 | 0.25 | 1.60 | 12 | 20/80 | 6/24 |
| 3.16 | 1.84 | +0.50 | 0.32 | 1.30 | 10 | 20/63 | 6/19 |
| 2.51 | 1.46 | +0.40 | 0.40 | 1.00 | 8 | 20/50 | 6/15 |
| 2.00 | 1.16 | +0.30 | 0.50 | 0.80 | 6 | 20/40 | 6/12 |
| 1.58 | 0.92 | +0.20 | 0.63 | 0.63 | 5 | 20/32 | 6/9.5 |
| 1.26 | 0.73 | +0.10 | 0.80 | 0.50 | 4 | 20/25 | 6/7.5 |
| 1.00 | 0.58 | 0.00 | 1.00 | 0.40 | 3 | 20/20 | 6/6.0 |
| 0.79 | 0.46 | -0.10 | 1.25 | 0.32 | 2 | 20/16 | 6/4.8 |
| 0.63 | 0.37 | -0.20 | 1.60 | 0.25 | 1.8 | 20/12.5 | 6/3.8 |
| 0.50 | 0.29 | -0.30 | 2.00 | 0.20 | 1.5 | 20/10 | 6/3.0 |

As the exact font used for each chart was not known, line spacing could not be accurately measured. However, as fonts were required to be similar to Helvetica or Times New Roman [1], the x-height ratio from these – the ratio of the x-height to full letter height – were extracted, and measured x-heights divided by this value to deduce the expected line height. Per the Python matplotlib package [12], this ratio is 0.523 for Helvetica (sans-serif) and 0.450 for Times New Roman (serif). Letters were separated into lines using the y-coordinates recorded for the bottom of each letter. Using the cap height calculated for each text size, letters were deemed to be on the first line of the passage if the y-coordinate deviated from the minimum recorded y-coordinate by less than the cap height and were deemed to be on the second line where this deviation was 1-2 times the cap height. The difference in mean y-coordinates for each line was calculated, then divided by the expected line height for that font type (serif or sans-serif) to find the line spacing. The overall line spacing for a given chart was calculated as the median line spacing for all font sizes. As this calculation relies on assumed font characteristics, line spacing was also calculated as the baseline-to-baseline measurement divided by the x-height, which is directly measurable and font agnostic.

Chart contrast was estimated using the scanned images cropped to include only the chart text. Contrast was calculated using the Weber formula as required by the standard:

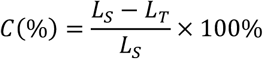

Where 𝐿_𝑆_ is the luminance of the surrounding field, and 𝐿_𝑇_ is the luminance of the text, with pixel intensity value used as a surrogate for luminance. Images were first converted to grayscale and gamma-corrected using the standard formula:

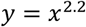

where 𝑥 is the pixel value normalised to the range [0,1]. Gaussian filtering with a kernel size of 9×9 pixels was then applied to account for text printed in halftone style and noise induced by the scanning process. 𝐿_𝑆_ was defined as the largest (brightest) pixel value and 𝐿_𝑆_ defined as the smallest (darkest) value. This contrast calculation is an estimate only and differs from the ISO requirement of contrast in standardised illumination and viewing distance conditions.

## Results

A total of 19 reading charts were collected and scanned. Chart 8 included two different charts, one on each side, which were measured separately as 8.1 and 8.2 and considered different charts. Charts 9 and 18 contained identical text with different designs so were also considered as different charts. Twelve charts (63%) used serif fonts, with the remaining seven using sans-serif (37%). Most cards used a mixture of notations permitted by the ISO standard (logMAR, Decimal, M size, N size, Snellen) as summarised in Table 2. N size was used exclusively in eight charts (42.1%). Three cards (16%) included Jaeger, which is a non-standardised unit [13], but also included other ISO-permissible units. The most common unit included was N size, present on 16 charts (84%), followed by logMAR on six charts (32%). No cards used all units permitted by the standard, and no cards used the RAD or logRAD terminology.

**Table 2:** Letter size units used for each chart. MAR is minimum angle of resolution. Jaeger, highlighted in grey, is not included in ISO 7921:2024 [1].

| Chart | Font serif | logMAR | Decimal | M size | N size | Snellen | Jaeger |
| --- | --- | --- | --- | --- | --- | --- | --- |
| 1 | Sans-serif |  | ✓ |  |  |  |  |
| 2 | Serif | ✓ |  |  | ✓ |  |  |
| 3 | Serif | ✓ |  |  | ✓ |  |  |
| 4 | Sans-serif |  |  |  | ✓ |  |  |
| 5 | Serif | ✓ |  |  | ✓ |  |  |
| 6 | Serif |  |  |  | ✓ |  |  |
| 7 | Sans-serif | ✓ | ✓ | ✓ |  | ✓ |  |
| 8.1 | Sans-serif | ✓ | ✓ |  | ✓ | ✓ | ✓ |
| 8.2 | Sans-serif | ✓ | ✓ |  | ✓ | ✓ | ✓ |
| 9 | Serif |  |  |  | ✓ |  |  |
| 10 | Serif |  |  |  | ✓ |  |  |
| 11 | Sans-serif |  |  |  | ✓ |  |  |
| 12 | Serif |  |  | ✓ | ✓ |  |  |
| 13 | Serif |  |  | ✓ | ✓ |  |  |
| 14 | Serif |  |  |  | ✓ |  |  |
| 15 | Serif |  |  |  | ✓ |  |  |
| 16 | Sans-serif |  | ✓ |  |  |  |  |
| 17 | Sans-serif |  |  | ✓ | ✓ | ✓ | ✓ |
| 18 | Serif |  |  |  | ✓ |  |  |

Text sizes included on each card are summarised in Table 3. The most common sizes were +0.20 and +0.30 logRAD (or equivalent), with both included in 18 charts (94.7%). Only four charts (21.1%) – charts 7, 12, 13 and 16 – contained 0.00 logRAD (or equivalent) as required by the standard. Three of these included size increments up to their maximum sizes as required by the standard.

**Table 3:** Text sizes included in each reading chart, converted to the nearest logRAD equivalent in accordance with ISO 7921:2024. Sizes marked with an asterisk (*) are included but mislabelled. Cells marked with a hash (#) were not a multiple of 0.10 log units so have been rounded to the nearest value. Underlined cells indicate an N or M size which was rounded to the nearest permissible value. Grey cells indicate x-heights derived from cap heights as there were no measurable lowercase letters. Orange cells indicate text with no measurable characters.

| Chart | logRAD |  |  |  |  |  |  |  |  |  |  |  |  |  |  |  |
| --- | --- | --- | --- | --- | --- | --- | --- | --- | --- | --- | --- | --- | --- | --- | --- | --- |
|  | 0.0 | 0.1 | 0.2 | 0.3 | 0.4 | 0.5 | 0.6 | 0.7 | 0.8 | 0.9 | 1.0 | 1.1 | 1.2 | 1.3 | 1.4 | 1.5 |
| 1 |  | ✓ | ✓ | ✓ | ✓ | ✓ | ✓ | ✓ |  |  | ✓ |  |  |  |  |  |
| 2 |  |  | ✓ | ✓ | ✓* |  | ✓ | ✓ |  | ✓ | ✓* |  |  | ✓* |  |  |
| 3 |  |  | ✓ | ✓ | ✓ |  | ✓ | ✓ |  | ✓ | ✓ |  |  | ✓ |  |  |
| 4 |  |  | ✓ | ✓ | ✓ | ✓ |  |  | ✓ |  |  |  |  |  |  |  |
| 5 |  |  | ✓ | ✓ | ✓ |  | ✓ | ✓ |  | ✓ | ✓ |  |  | ✓ |  |  |
| 6 |  |  | ✓ |  | ✓ |  | ✓ | ✓ |  | ✓ | ✓ | ✓ | ✓ |  |  | ✓ |
| 7 | ✓ | ✓ | ✓ | ✓ | ✓ | ✓ | ✓ | ✓ |  |  |  |  |  |  |  |  |
| 8.1 |  | ✓ | ✓ | ✓ | ✓ | ✓ |  | ✓ |  |  | ✓# |  |  |  |  |  |
| 8.2 |  | ✓ | ✓ | ✓ |  | ✓ | ✓ | ✓ |  |  | ✓# |  |  |  |  |  |
| 9 |  | ✓ | ✓ | ✓ | ✓ | ✓ | ✓ | ✓ | ✓ | ✓ |  | ✓ |  |  |  |  |
| 10 |  | ✓ | ✓ | ✓ | ✓ | ✓ | ✓ | ✓ | ✓ | ✓ |  | ✓ | ✓ |  |  |  |
| 11 |  |  | ✓ | ✓ | ✓ | ✓ |  | ✓ | ✓ |  | ✓ |  |  |  |  |  |
| 12 | ✓ | ✓ | ✓ | ✓ | ✓ | ✓ |  |  |  |  |  |  |  |  |  |  |
| 13 | ✓ | ✓ | ✓ | ✓ | ✓ | ✓ |  |  |  |  |  |  |  |  |  |  |
| 14 |  |  | ✓ | ✓ | ✓ | ✓ | ✓ |  |  |  |  |  |  |  |  |  |
| 15 |  |  |  | ✓ |  |  |  |  |  |  |  |  |  |  |  |  |
| 16 | ✓ | ✓ | ✓ | ✓ | ✓ | ✓ |  | ✓ |  |  | ✓ |  |  |  |  |  |
| 17 |  | ✓ | ✓ | ✓ | ✓ | ✓ |  | ✓ |  |  | ✓ |  |  |  |  |  |
| 18 |  | ✓ | ✓ | ✓ | ✓ | ✓ | ✓ | ✓ | ✓ | ✓ |  | ✓ |  |  |  |  |

### Labels

Labelling inconsistencies were evident on some charts. Many charts provided multiple labels for each text size, though some of these were not in agreement with the conversion table in the ISO standard. Other charts contained clear labelling errors. These findings are annotated in Table 3.

Chart 2 contained N size and logMAR labels, with inconsistencies for +0.40, +1.00 and +1.30 logMAR. The chart included duplicate +0.70 and +1.30 logMAR lines which corresponded to incorrect N sizes. Charts 2, 3, 5, 6, 11, 16 and 17 contained N or M sizes which were not included in the standard, and did not include other units, so were rounded to the nearest permissible value. Charts 12, 13 and 17 provided decimal notation but did not note the viewing distance, and charts 4, 6, 9, 10, 11, 15 and 18 only provided N size without a viewing distance. In these cases, a 40cm viewing distance was assumed for the purposes of comparison against the standard.

Charts 8.1 and 8.2 contain Snellen notations at 40cm but have used an incorrect conversion. For example, N4 (+0.10 logMAR) should correspond to a reduced Snellen fraction of 20/25 but is listed as 40/25, which has then been converted to a decimal value of 1.60. These Snellen and decimal values correspond to N1.8 or −0.20 logMAR which is much smaller than the printed text. If assuming the numerators should be 20 in the recorded Snellen fractions, N10 (40/70) and N26 (40/200) are still incorrect. As a result, the logRAD conversion from N size has been used instead of the listed logMAR sizes.

### Text size

From the 19 charts, a total of 133 text sizes were measured by each of the three graders (total 399). Inter-grader agreement of x-height measurements ICC(2,1) was excellent at 0.99989 (95% CI 0.99961-0.99995, F(124,248)=54557.60, p<0.001). Note that this score has been presented to five decimal places here due to the unusual nature of the result, but will be rounded to two decimal places for the remainder of the manuscript.

### Serif charts

None of the 12 serif charts were within tolerance for all sizes (Fig. 3), and all texts larger than +0.70 logRAD were too small. Chart 14 had letters too large for +0.20 logRAD, with no other charts having text that were larger than labelled. Overall, sans-serif charts were smaller (µ=−9.63%, σ=8.53, range [−29.80, 18.31]), resulting in an underestimation of near acuity.

**Fig. 3:**
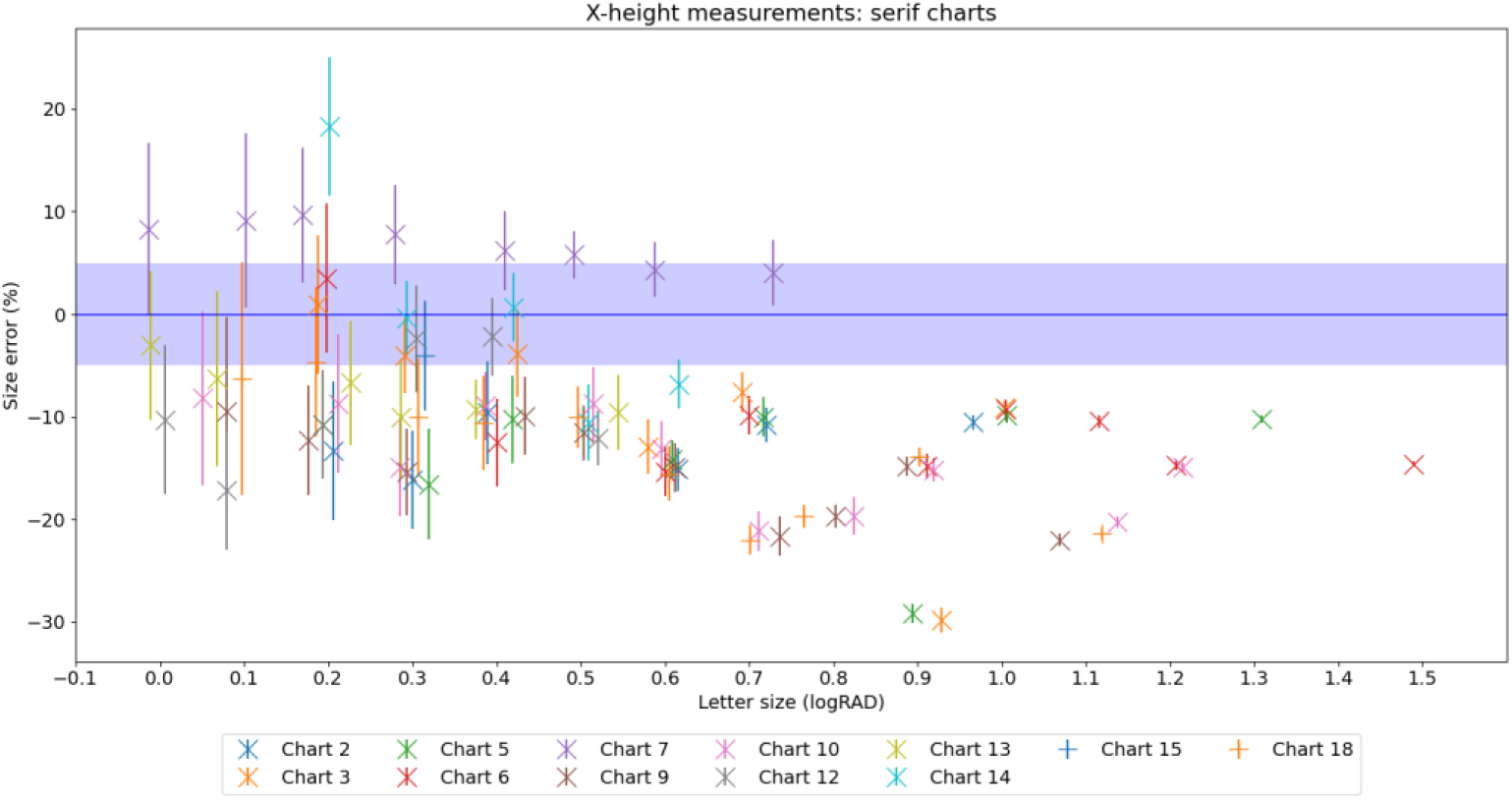
Differences in measured font x-heights for serif charts compared to the expected value (dark blue line) and tolerance (light blue bar). Error bars indicate the measurement error of the pixel counting method and mean grader standard deviation for that chart size. Horizontal jitter has been added for readability.

### Sans-serif charts

Of the seven sans-serif font charts, only chart 17 was within tolerance for all font sizes. Deviations from the expected sizes are shown in Fig. 4. Chart 1 was consistently larger than the expected size. Charts 8.1 and 8.2 showed smaller text measuring too big and larger text measuring too small. Charts 4, 11 and 16 were in tolerance for most sizes. Overall, sans-serif charts did not significantly differ from the standard (µ=+4.96%, σ=9.78, range [−16.11, 23.95]).

**Fig. 4:**
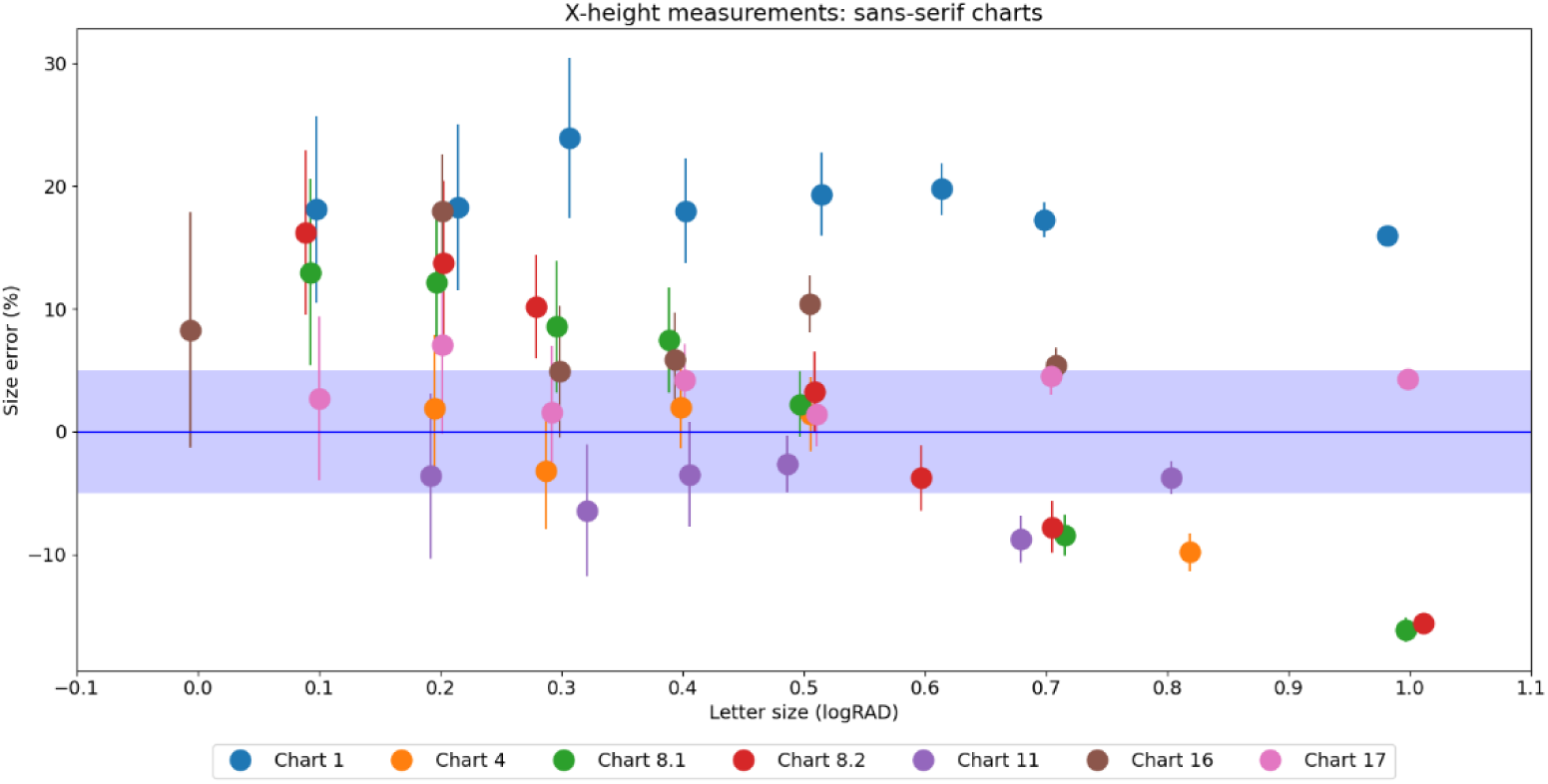
Differences in measured font x-heights for sans-serif charts compared to the expected value (dark blue line) and tolerance (light blue bar). Error bars indicate the measurement error of the pixel counting method and mean grader standard deviation for that chart size. Horizontal jitter has been added for readability.

Comparing the measurements to the rounded N sizes instead of expected x-height results in the same size error for serif (µ=+8.91%, σ=9.57) and sans-serif (µ=+8.40%, σ=9.72) (U=2091.0, p=0.67), as shown in Fig. 5.

**Fig. 5:**
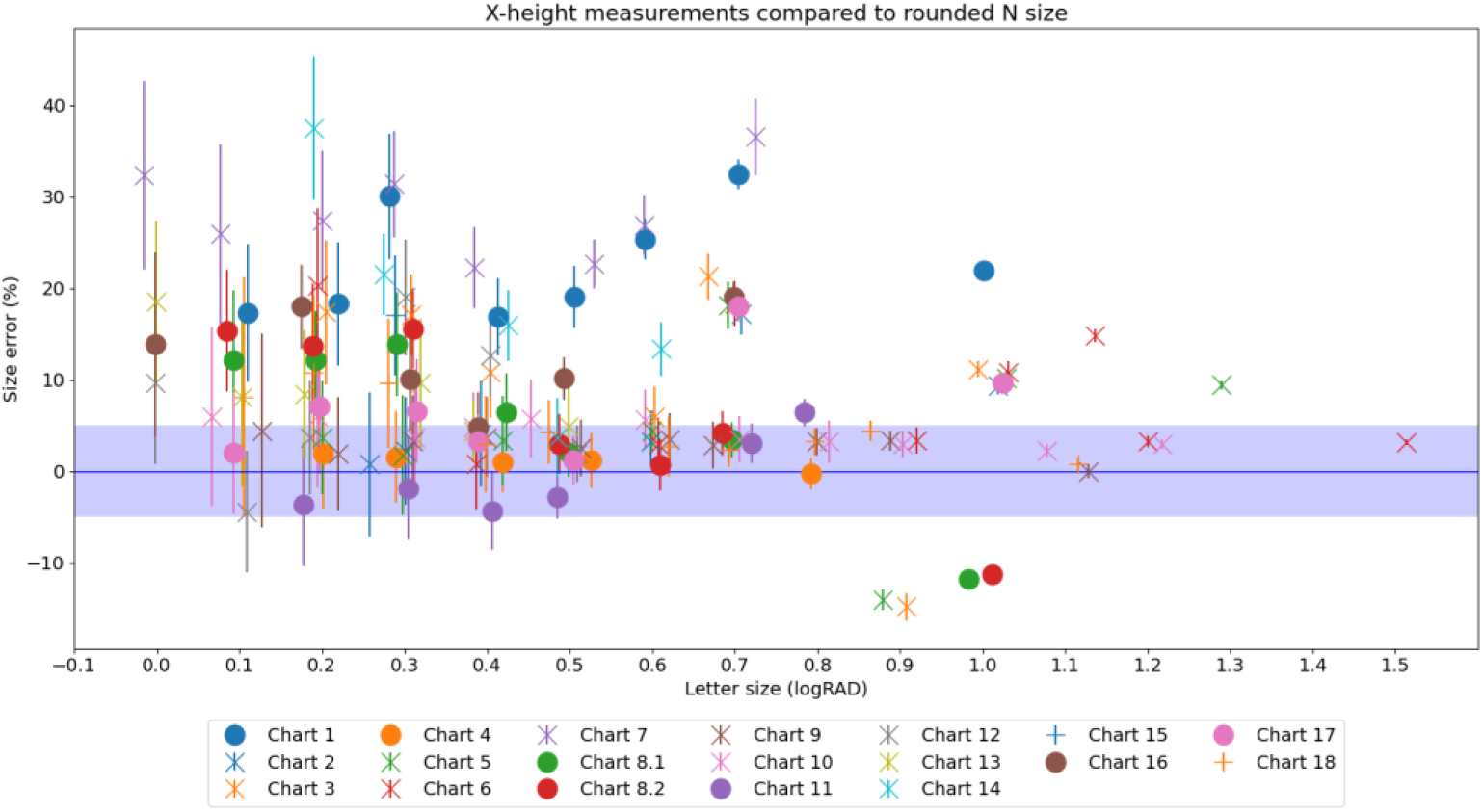
Differences in measured font heights for near acuity charts compared to the rounded N sizes (dark blue line) and tolerance (light blue bar). Error bars indicate the measurement error of the pixel counting method and mean grader standard deviation for that chart size. Round markers indicate sans-serif fonts with crosses indicating serif fonts. Horizontal jitter has been added for readability.

Calculating expected x-heights from font sizes relies on the assumed x-height ratios being correct, however, due to the estimation process used in this study, these results may be inaccurate.

### Line spacing

The line spacing values for each chart are summarised in Table 4, along with baseline-to-baseline:x-height ratios. The standard requires spacing to be less than or equal to 1.5, which was observed for all charts. Notably, the calculated line spacing varied markedly between lines, as reflected by the minimum and maximum line spacing values. Analysis of the scanned images agreed with this variance, suggesting different line spacings for different text sizes. Where variability was noted, the larger text sizes tended to have smaller line spacing. Charts 6, 7 and 14 had median line spacings less than 1.0, with all other charts between 1.0 and 1.3. While there is no standard for baseline-to-baseline:x-height ratio, charts overall had a ratio of 2.35 (IQR 0.36).

**Table 4:**
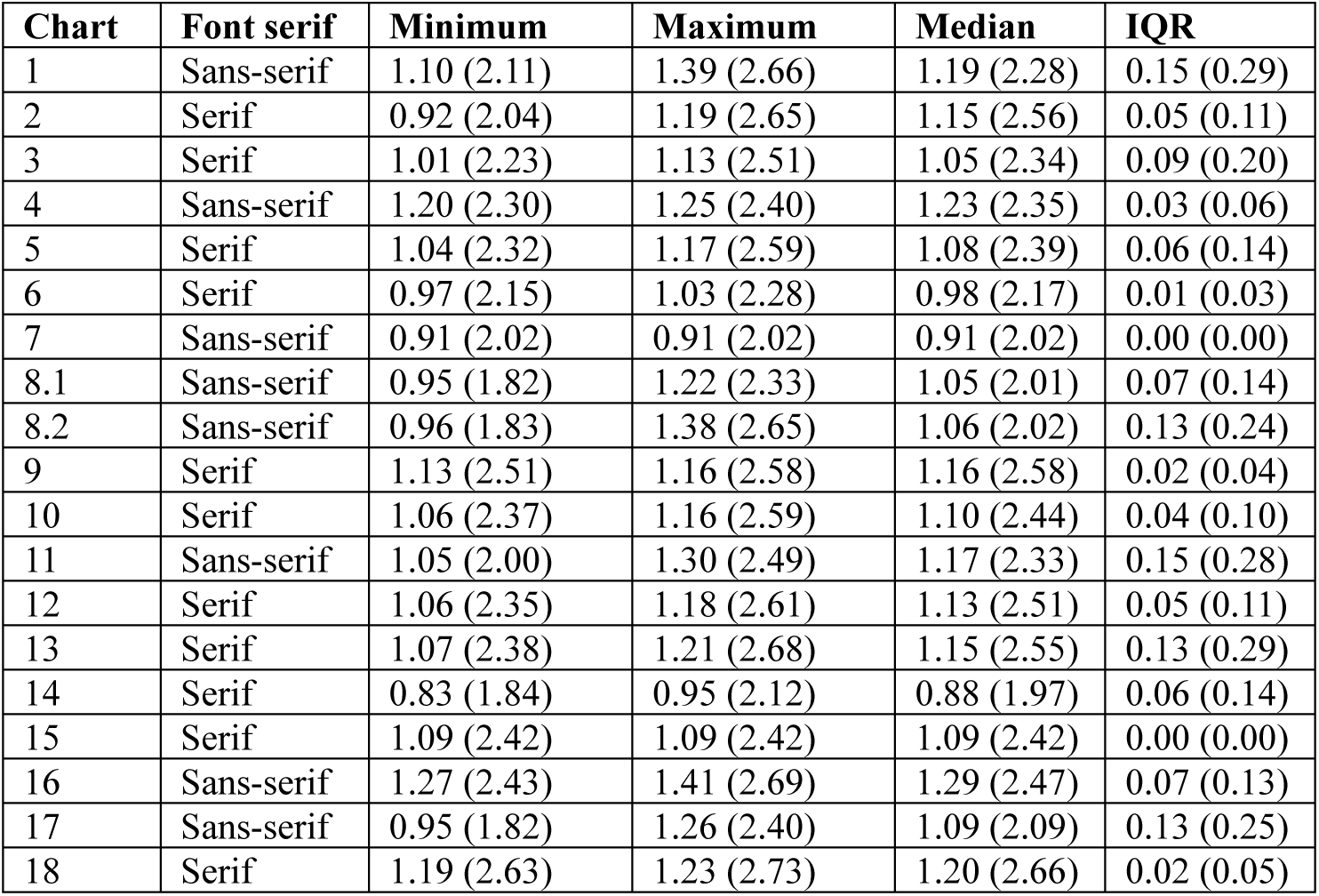
Estimated line spacing for each chart. Values are calculated based on the x-height ratios of Helvetica (sans-serif) or Times New Roman (serif). Values in brackets are baseline-to-baseline:x-height ratios.

### Contrast

Weber contrast was estimated for all charts. All charts had a white background (pixel value 255) with text values ranging from 19 to 94. The lowest contrast value was 88.9% for chart 17, which exceeded the minimum 85.0%. The standard requires text be printed on a matte surface to minimise specular reflections as this can further reduce contrast [1], and matte surfaces were used for 11 (58%) of the charts. A full breakdown of these data are shown in Table 5, along with each chart’s country of origin.

**Table 5:**
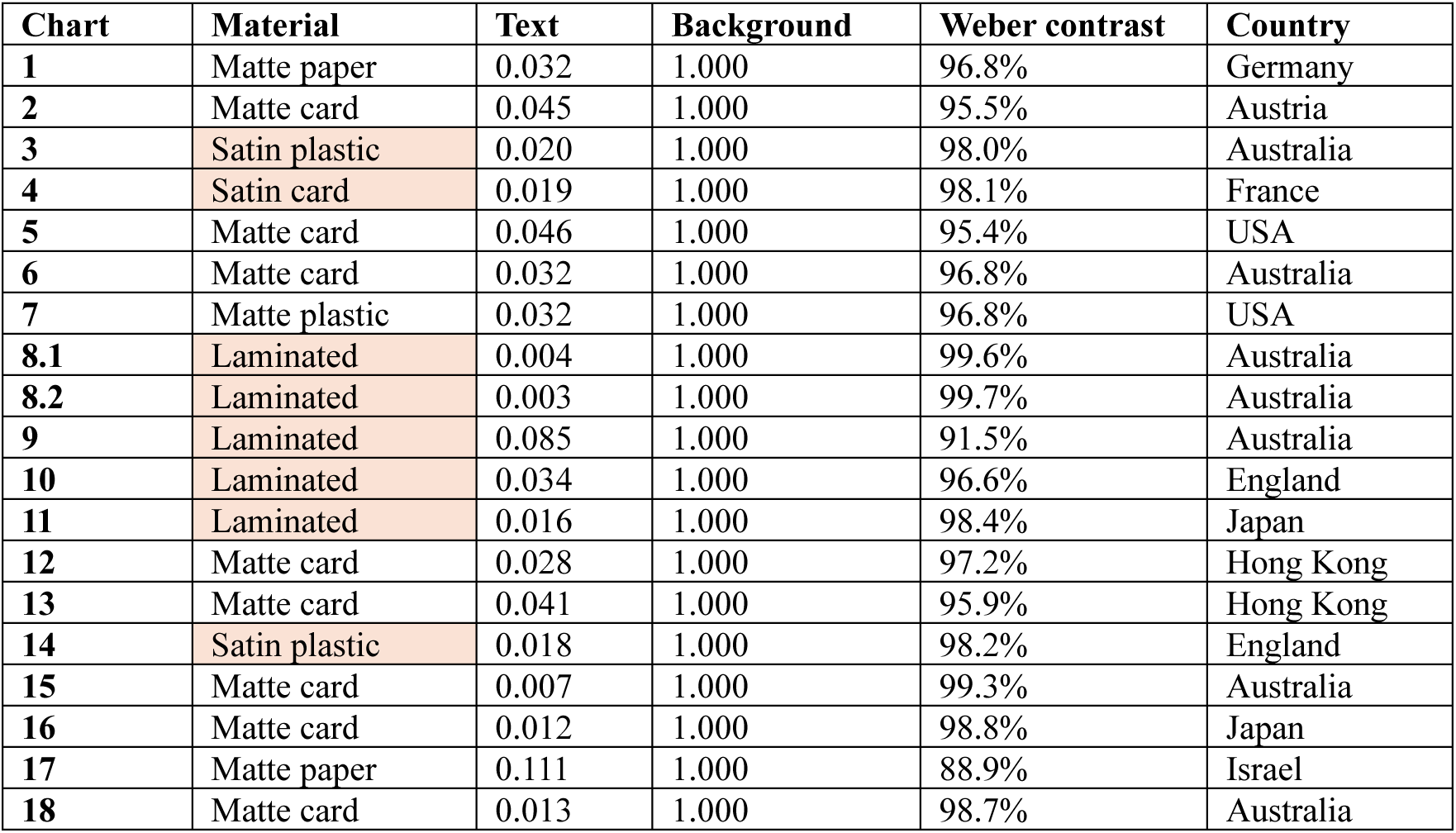
Weber contrast, chart material and manufacturer country details for each chart. Text and background values are pixel intensities in the range [0,1] and rounded to three decimal places. Highlighted cells indicate contrast less than 85% or a non-matte surface, both of which are inconsistent with ISO 7921:2024 [1].

| Chart | Material | Text | Background | Weber contrast | Country |
| --- | --- | --- | --- | --- | --- |
| 1 | Matte paper | 0.032 | 1.000 | 96.8% | Germany |
| 2 | Matte card | 0.045 | 1.000 | 95.5% | Austria |
| 3 | Satin plastic | 0.020 | 1.000 | 98.0% | Australia |
| 4 | Satin card | 0.019 | 1.000 | 98.1% | France |
| 5 | Matte card | 0.046 | 1.000 | 95.4% | USA |
| 6 | Matte card | 0.032 | 1.000 | 96.8% | Australia |
| 7 | Matte plastic | 0.032 | 1.000 | 96.8% | USA |
| 8.1 | Laminated | 0.004 | 1.000 | 99.6% | Australia |
| 8.2 | Laminated | 0.003 | 1.000 | 99.7% | Australia |
| 9 | Laminated | 0.085 | 1.000 | 91.5% | Australia |
| 10 | Laminated | 0.034 | 1.000 | 96.6% | England |
| 11 | Laminated | 0.016 | 1.000 | 98.4% | Japan |
| 12 | Matte card | 0.028 | 1.000 | 97.2% | Hong Kong |
| 13 | Matte card | 0.041 | 1.000 | 95.9% | Hong Kong |
| 14 | Satin plastic | 0.018 | 1.000 | 98.2% | England |
| 15 | Matte card | 0.007 | 1.000 | 99.3% | Australia |
| 16 | Matte card | 0.012 | 1.000 | 98.8% | Japan |
| 17 | Matte paper | 0.111 | 1.000 | 88.9% | Israel |
| 18 | Matte card | 0.013 | 1.000 | 98.7% | Australia |

## Discussion

None of the tested reading charts met all requirements of the ISO 7921:2024 standard. Interestingly, the only chart to meet the text size requirements was a free chart provided by a lens manufacturer, though this chart did have inconsistencies with labelling. Although none of the charts claim conformance with this standard, the variability in charts analysed in this study, and lack of standards compliance, is of clinical significance. Conditions including age-related macular degeneration, cataracts and presbyopia rely on changes in near reading acuity for effective management [8], and the significant variability in charts may mask or exaggerate subtle changes.

In Australia and the United Kingdom, reading acuity is commonly recorded using N size as it is an easily understood metric for patients based on a point system similar to the labelling of font sizes on the computer [4, 14]. However, the conversion table provided in the ISO standard notes that the listed N sizes are approximations (does not follow a logarithmic progression for letters larger than N12), rounded to the nearest common size based on the properties of the Helvetica font, and should only be used to identify the near reading acuity grade [1]. As the reading acuity angle is based on the font’s x-height, the true font size depends on the ratio of x-height to font height. For Helvetica, this ratio is 0.523 according to the font file included in the matplotlib python package [12], meaning a 10pt font would have an x-height of 5.23pt. The standard notes that serif fonts shall be similar to Times New Roman, which has a height to x-height ratio of 0.450 [12]. Thus, serif fonts need to be 1.16 times the size of their sans-serif counterparts to have matching x-heights, as shown in Table 6. As each 0.10 log unit step provides a 26% increase, this results in a given size in Times New Roman being closer to the next largest size in Helvetica.

**Table 6:** Actual font sizes for Helvetica and Times New Roman (Times) for each logRAD value, compared to the rounded sizes provided in the standard (ISO). Values are rounded to 3 significant figures.

| logRAD | N size (ISO) | Points (Helvetica) | Points (Times) |
| --- | --- | --- | --- |
| +1.30 | 60 | 62.8 | 73.1 |
| +1.20 | 48 | 49.9 | 58.1 |
| +1.10 | 36 | 39.6 | 46.1 |
| +1.00 | 30 | 31.5 | 36.7 |
| +0.90 | 24 | 25.0 | 29.1 |
| +0.80 | 18 | 19.9 | 23.1 |
| +0.70 | 14 | 15.8 | 18.4 |
| +0.60 | 12 | 12.5 | 14.6 |
| +0.50 | 10 | 9.95 | 11.6 |
| +0.40 | 8 | 7.91 | 9.21 |
| +0.30 | 6 | 6.28 | 7.31 |
| +0.20 | 5 | 4.99 | 5.81 |
| +0.10 | 4 | 3.96 | 4.61 |
| 0.00 | 3 | 3.15 | 3.66 |
| -0.10 | 2 | 2.50 | 2.91 |
| -0.20 | 1.8 | 1.99 | 2.31 |
| -0.30 | 1.5 | 1.58 | 1.84 |

Our measurements found the majority of charts to be outside of tolerance for font size, with serif fonts generally too small. This matches the differences in equivalent point sizes for serif and sans-serif fonts if charts are printed using rounded N sizes instead of x-heights; 4-point font will be approximately correct for Helvetica (3.96) but too small for Times New Roman (4.61). The equivalent size errors of serif and sans-serif charts when comparing x-heights to the expected x-heights at a given point size (Fig. 5) suggests manufacturers are using the N size as a surrogate for font size (in points) without considering x-heights.

Estimated line spacing was found to meet the requirements of the standard; however, the spacing differed between text sizes on some charts. It is speculated that this may be a design decision, with larger text printed with smaller line spacing to fit more text on the page. While the standard does not require consistent spacing to be used throughout the chart, this variation may have an effect on crowding, which has been shown to affect visual acuity in healthy and diseased retinas [15, 16]. Again, these calculations relied on assumed x-height ratios, so this may not be accurate.

When considering the baseline-to-baseline:x-height ratio, line spacing can be directly compared to the font’s x-height and removes ambiguity around the font’s size and spacing above the ascender. For Times New Roman the ascender is located at 0.683 times the full letter height (as measured from baseline) compared to 0.718 for Helvetica [12], meaning the top 31.7% of the Times New Roman text is spacing compared to 28.2% for Helvetica. With Times New Roman font needing to be 16% larger than Helvetica to match x-heights, the spacing above the ascender for Times New Roman becomes 30% larger than Helvetica. Indeed, Table 4 shows smaller line spacing values for serif fonts compared to sans-serif for similar baseline-to-baseline:x-height values. This disparity, combined with the inability to directly measure line spacing without knowledge of the font’s properties, calls into question the utility of line spacing in the ISO standard. Further work is warranted to determine whether a more objective and measurable metric, such as the baseline-to-baseline:x-height ratio, is more appropriate.

Interestingly, most charts did not include 0.00 logRAD (or equivalent) and many charts measured in this study provided +0.10 logRAD (or equivalent) as the smallest size. Although +0.10 logRAD is not commonly encountered in the real-world, including smaller sizes in reading acuity charts may improve the ability to detect small changes in vision. Furthermore, Radner and Benesch [17] found near acuity to be better than 0.00 logRAD in healthy patients between 25 and 74 years of age, and around −0.10 logRAD between 25 and 54, so limiting testing to +0.10 logRAD may not allow changes to be detected until near acuity has dropped by more than two lines. Indeed, distance acuity is routinely measured to thresholds at or below 0.00 logMAR for this reason.

All charts tested in this study exceeded the minimum required contrast as measured using gamma-corrected scans. It is important to note that this technique is different from directly measuring luminance from the physical chart when the background luminance is in the range 80-200 cd/m^2^ as described in the standard. This digital surrogate should be considered an estimation. Since all charts were black or dark grey text on a white background, the authors are confident that the charts meet the minimum contrast requirements.

Using a scanner to measure two-dimensional objects has been validated as an accurate technique [18], though to the authors’ knowledge, this technique has not been applied to text measurements. A previous study by Chen et al. [19] analysed text size using a surgical microscope at 10x magnification, though no repeatability statistics are available. Our technique is highly repeatable, as evidenced by the ICC score of 1.00, and provides a mechanism for verification by third parties by assessing the same types of images. Our testing found the method to be highly accurate, which may be due to the simple black-and-white nature of reading charts, and elimination of grey pixels. Additional validation of this technique using other printed materials with varying contrast and chromaticity is warranted.

The difference between the smallest and largest measurement was greater than 30% for all sizes between +0.10 and +0.70 logRAD, which is greater than the 0.10 log unit (26%) progression. This chart variability reinforces the need to use standardised equipment, especially in disease monitoring and research. Clinics should endeavour to use charts which conform to ISO 7921:2024 or, where this is not possible, ensure the same chart is used throughout the practice and across visits. Near visual acuity recording should be accompanied by both the chart name and test distance to ensure repeatability. Shared care arrangements should ensure all practitioners are using ISO compliant charts, or at a minimum, the same near chart, for monitoring and progression.

These results prompted the authors to create a new chart, the UC/UWA Reading Chart, which fully conforms to ISO 7921:2024. This chart, available in the supplementary material, includes text sizes from −0.10 to +0.90 logRAD in 0.10 steps and is available in both serif and sans-serif versions. Both versions have been validated by converting the chart to an image at 2540 dpi, providing 0.01mm granularity as required by the standard, and measuring the heights of ‘x’ characters from −0.30 to +1.30, with all sizes within tolerance. These charts have been made available at https://github.com/tim-murphy/near_chart under the Creative Commons BY-ND 4.0 license (https://creativecommons.org/licenses/by-nd/4.0/) to provide access to researchers and clinicians without financial constraints or use restrictions.

While the charts are freely available in digital form, charts must be printed and validated for conformance with the ISO standard via a procedure detailed in the chart’s accompanying documentation. To ensure text is legible and within size tolerances, the chart must be printed at a resolution of 1200 dpi or higher to ensure the printing size error is within 5% of the required values. The text used on the charts are not linguistically standardised and should not be used to compare reading speeds or for any other purposes requiring text standardisation. Further information is available in the supplementary materials.

### Intraclass correlation

The exceptionally high ICC score of 1.00 is surprising and requires further explanation. This result suggests the three graders had almost perfect agreement across all letter sizes for all charts. While this seems unusual at face value, adaptive thresholding was undertaken before letters were measured to minimise subjectivity, thereby maximising agreement. As illustrated in Fig. 2, the y-coordinate of the pixels at the top and bottom of the ‘w’ is clearly defined so perfect agreement is expected for this case. Adaptive thresholding is not perfect, and scanned materials are slightly slanted, so small measurement differences were noted between letters on the same line.

However, these differences were no larger than two pixels when considering all letters measured by all graders for a given chart and text size. Furthermore, the mean measurement was used for each grader for each chart and text size which further reduced intergrader disparity. Raw data have been included as supplementary material for transparency.

### Limitations

This study is not without limitations. Measurements were undertaken using a 600 dpi scanner, which has a granularity of 0.0423mm. The standard requires verification to be undertaken with devices having a measurement error of 0.01mm or less. Such accuracy would require a scanner with resolution of 2540 dpi or greater, which are not widely available. As this study has a larger measurement error, text may have been labelled as within tolerance when they were not. Most charts were found to be outside of tolerance for at least one size, and this limitation does not invalidate any findings of non-conformance, though it may suggest charts are closer to the standard than they are. The practicality of measuring at 0.01mm is challenging due to ink bleed and other printing imperfections, and it is not clear whether the same technique would work at such a high resolution. Future work using higher resolution methods would be beneficial.

The fonts used for each chart cannot be extracted as the manufacturers do not publish this information. While this does not affect the x-height measurements, it limits the ability to infer x-height from cap height where no appropriate letters were printed on the chart. This also creates uncertainty when assessing line spacing. As a result, results relying on x-height ratios for cap height should be read with caution.

Finally, this study used charts which were available to the researchers. This is a limited subset and may not be representative of charts found in clinics globally. The authors encourage replication of this work using other charts and have made the software available at https://github.com/tim-murphy/near_chart.

## Conclusion

Near reading acuity is commonly measured in clinical practice and in research. This research highlighted the variability between commercially-available and ophthalmic device manufacturer reading charts, with none of the available charts meeting all requirements of the ISO 7921:2024 standard. As such, clinicians should be cautious when comparing acuity results using different reading charts, especially when monitoring progressive diseases or as part of shared-care models. Future research should explore whether the differences between charts are clinically significant. A new vision chart was proposed by the authors in response to the findings of the study.

## Supporting information

threshold verification

data for ICC statistics

UC/UWA Reading Chart v0.91

## Data Availability

Software used for analysis is available at https://github.com/tim-murphy/near_chart.
The data that support the findings of this study are not openly available due to copyright restrictions and are available from the corresponding author upon reasonable request. Data are located in controlled access data storage at the University of Canberra.

## Acknowledgements

The authors would like to thank the staff in the Discipline of Optometry and Vision Science at the University of Canberra for their feedback on the development of the UC/UWA Reading Chart.

## Declarations

### Data availability statement

Software used for analysis is available at https://github.com/tim-murphy/near_chart.

The data that support the findings of this study are not openly available due to copyright restrictions and are available from the corresponding author upon reasonable request. Data are located in controlled access data storage at the University of Canberra.

### Author contributions

Conceptualisation: Timothy Murphy; Methodology: Timothy Murphy, Jingyi Chen, Myra Leung; Formal analysis and investigation: Timothy Murphy, Jingyi Chen, Myra Leung; Writing – original draft preparation: Timothy Murphy, Jingyi Chen, Myra Leung; Writing – review and editing: Timothy Murphy, Jingyi Chen, Myra Leung; Resources: Timothy Murphy; Software: Timothy Murphy

