## Supplementary material for "How accurate are our near reading charts? An assessment of 19 charts against ISO standards": threshold verification

### Threshold measurement verification

The image below shows the scanned ruler with thresholding applied. The image below shows the ruler scanned in a vertical orientation and rotated 90 degrees to display horizontally. Pixel measurements were taken from the top-right corner of the zero marker (256, 122) and top-right corner of the 20cm marker (4980, 184). To account for scans being slightly slanted, pixel distance was calculated using Pythagoras' theorem as 4724.4 pixels. The expected pixel count at 600dpi is 4724.4 pixels.

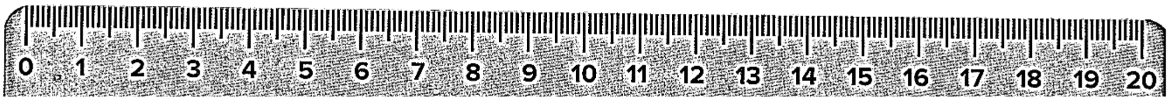


The same technique was repeated with the ruler in a horizontal orientation. Pixel measurements were taken from the bottom-right corner of the zero (124, 225) and 20cm markers (4854, 232), providing a pixel distance of 4730.0 pixels This resulted in an error of 0.12%.

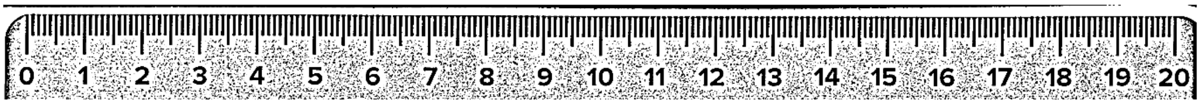
