## Supplementary material for "How accurate are our near reading charts? An assessment of 19 charts against ISO standards": UC/UWA Reading Chart v0.91

| +0.90 logRAD | 6/48 | 0.125 | 3.2M | N24 |
|---|---|---|---|---|

Computer reading glasses are designed to reduce eye strain and keep things clear when doing screen work.

zones — vision — woven

| +0.80 logRAD | 6/38 | 0.16 | 2.5M | N18 |
|---|---|---|---|---|

Bifocal glasses have two zones: the top part for far away and the bottom part for reading. They allow clear vision at different distances.

screen — waxes — axons

| +0.70 logRAD | 6/30 | 0.20 | 2.0M | N14 |
|---|---|---|---|---|

Long periods of near work can cause headaches, tired eyes, and make it harder to keep things in focus. Regular breaks and good lighting can help reduce strain and improve comfort.

aroma — oceans — rescue

| +0.60 logRAD | 6/24 | 0.25 | 1.6M | N12 |
|---|---|---|---|---|

The brain and eyes work together to allow people to see. Although each eye sees a slightly different view, the brain combines the two views together to form one image.

nearer — sesame — wanes

**UC/UWA Reading Chart**

Version 0.91.serif (April 2026)

To be used at 40cm

Manufactured by the University of Canberra, Bruce ACT 2617, Australia

Murphy TI, Chen J, Leung M. How accurate are our near reading charts? An assessment of 19 charts against ISO standards. medrxiv. 2026 Jan 30:2026-01.

| +0.50 logRAD | 6/19 | 0.32 | 1.3M | N10 |
|---|---|---|---|---|

It is common for some people to see floaters, which can appear as dark or light spots or strings that move with the eye. If there are sudden increases in floaters, or flashing lights, this could indicate damage to the retina at the back of the eye which needs urgent attention.

sermon — common — assume

| +0.40 logRAD | 6/15 | 0.40 | 1.0M | N8 |
|---|---|---|---|---|

For some people the front surface of the eye or the lens inside the eye has an uneven shape. This is called astigmatism and can cause people to see ghost images or shadows around objects up close and far away. Glasses or contact lenses can usually help make things clearer and reduce eye strain.

answer — secure — casino

| +0.30 logRAD | 6/12 | 0.50 | 0.80M | N6 |
|---|---|---|---|---|

When the eye is too short it has to work harder to see things, especially up close. This is called long-sightedness or hyperopia, and can be tiring as the eye has to work harder than normal to see things clearly. Some people also get headaches after long periods of reading or screen time. Glasses can help to adjust the eye's focus, making close-up tasks like reading and sewing more comfortable.

ensure — sauna — waves

| +0.20 logRAD | 6/9.5 | 0.63 | 0.63M | N5 |
|---|---|---|---|---|

People who can see clearly up close but not far away have short-sightedness. This is also known as myopia and is usually caused by the eyeball being too long. It normally develops in childhood and can worsen into early adulthood. The risk of eye problems increases as the eye grows, so it is important to manage myopia early. This can involve special contact lenses, glasses, or eyedrops.

arena — seasons — owner

| +0.10 logRAD | 6/7.5 | 0.80 | 0.50M | N4 |
|---|---|---|---|---|

Inside the eye, there is a lens that helps focus light so that a person can see. A cataract is a normal change where the lens slowly becomes cloudy with age. Cataracts can cause vision to appear hazy and colours to appear less vibrant. In the early stages it can be managed with better lighting, but will need surgery once vision and quality of life are significant affected.

cannon — worms — arrow

| 0.00 logRAD | 6/6.0 | 1.0 | 0.40M | N3 |
|---|---|---|---|---|

As we get older, it becomes harder for the lens inside the eye to change shape, which makes reading small print more difficult. This is a normal change known as presbyopia and is not a sign of disease. Many people first notice it when holding a book or phone a little farther away than usual. Reading glasses or multifocal lenses can make near tasks clearer and more comfortable, with most people continuing their daily activities without any significant issues.

source — cursor — ransom

| –0.10 logRAD | 6/4.8 | 1.3 | 0.32M | N2 |
|---|---|---|---|---|

The inside of the eye is lined with a layer of cells called the retina. These cells respond to light and enable us to see. As we get older these cells become less efficient, making it harder to adapt when moving from bright sunlight into a dark room. This is a natural change but can be quite frustrating when it affects everyday activities. Good lighting can make it easier for the eyes to adjust.

camera — reason — noise

UNIVERSITY OF CANBERRA

Conforms to ISO 7921:2024 when validated according to the accompanying documentation.

Validated by: ______________________

Signed: ______________________ Date: __________

THE UNIVERSITY OF WESTERN AUSTRALIA

### UC/UWA Reading Chart

#### Version 0.91.serif (April 2026)

This reading chart conforms to ISO 7921:2024 provided the following checks are performed. It is the user's responsibility to ensure this has been completed in full before use in a clinical or research setting. The manufacturers make no guarantee of the accuracy of this reading chart, or conformance to ISO 7921:2024, should this procedure not be followed. Once all checks listed below have been completed, document that the chart has been validated by dating and signing the bottom of the back page in the space provided.

**Printing**
To ensure all text is printed legibly and in the correct size, this chart must be printed at a mimimum 1200 dpi on white, A4 paper or cardboard. This chart should have a matte surface to avoid reflections which may impact contrast. No additional margins or padding can be added as this will scale the text. This chart may be printed in colour or grayscale. Check legibility of the smaller sizes with a magnifying device.

**Text size**
The PDF version of this chart contains text of the correct size, within the margin of error allowed by the standard. To ensure accuracy, a validation page is printed on the reverse of this page. The rulers in the margins should be accurate and can be checked with a ruler. Each line displays the logRAD size with three 'x' characters on either side. The height of these characters should be within 5% of the corresponding x-height in the table below. Measurements must be done using a device with a measurement error of 0.01mm or less, such as callipers.

**Contrast**
Text must have a contrast of at least 85%. The PDF version of this chart has a 100% contrast but this may be reduced by the printing process. Contrast is calculated by measuring the luminance of the background surrounding the text ($L_b$) and luminance of the text itself ($L_t$). Background luminance must be between 80 and 200 cd/m$^2$ at a distance of 40cm (80-120 cd/m$^2$ recommended). The chart contrast is calculated as $C = \frac{L_b - L_t}{L_b}$. If $C \geq 0.85$ then the chart has sufficient contrast.

| **Reading acuity angle** min of arc | **x-height** mm | **Near reading acuity grades at 40cm** | | | | | |
|---|---|---|---|---|---|---|---|
| | | **logRAD**[a] | **Decimal reading acuity**[a,b] | **M size**[a] | **N size**[c] | **Reduced Snellen fraction** | |
| 20.0 | 11.61 | +1.30 | 0.05 | 8.00 | 60 | 20/400 | 6/120 |
| 15.8 | 9.22 | +1.20 | 0.063 (0.06) | 6.30 | 48 | 20/320 | 6/95 |
| 12.6 | 7.33 | +1.10 | 0.08 | 5.00 | 36 | 20/250 | 6/75 |
| 10.0 | 5.82 | +1.00 | 0.10 | 4.00 | 30 | 20/200 | 6/60 |
| 7.94 | 4.62 | +0.90 | 0.125 | 3.20 | 24 | 20/160 | 6/48 |
| 6.31 | 3.67 | +0.80 | 0.16 | 2.50 | 18 | 20/126 | 6/38 |
| 5.01 | 2.92 | +0.70 | 0.20 | 2.00 | 14 | 20/100 | 6/30 |
| 3.98 | 2.32 | +0.60 | 0.25 | 1.60 | 12 | 20/80 | 6/24 |
| 3.16 | 1.84 | +0.50 | 0.32 (0.3) | 1.30 | 10 | 20/63 | 6/19 |
| 2.51 | 1.46 | +0.40 | 0.40 | 1.00 | 8 | 20/50 | 6/15 |
| 2.00 | 1.16 | +0.30 | 0.50 | 0.80 | 6 | 20/40 | 6/12 |
| 1.58 | 0.922 | +0.20 | 0.63 (0.6) | 0.63 | 5 | 20/32 | 6/9.5 |
| 1.26 | 0.733 | +0.10 | 0.80 | 0.50 | 4 | 20/25 | 6/7.5 |
| 1.00 | 0.582 | 0.00 | 1.00 | 0.40 | 3 | 20/20 | 6/6.0 |
| 0.79 | 0.462 | −0.10 | 1.25 | 0.32 | 2 | 20/16 | 6/4.8 |
| 0.63 | 0.367 | −0.20 | 1.60 | 0.25 | 1.8 | 20/12.5 | 6/3.8 |
| 0.50 | 0.292 | −0.30 | 2.00 | 0.20 | 1.5 | 20/10 | 6/3.0 |

[a] Values that end in zero may be truncated to delete the last zero only for the purpose of identifying the near reading acuity grade.
[b] Values in parentheses shall be used only for the purpose of identifying the near reading acuity grade.
[c] Values are approximations for Helvetica and rounded to the nearest common size for the purpose of identifying the near reading acuity grade.

**Use and care instructions**
This chart is to be used at 40cm under ambient or task lighting. Light sources not intended to illuminate the chart should not create specular reflections, and light sources visible to the subject may not exceed the chart background luminance. To ensure the print quality of this reading chart, store away from direct sunlight. The validation instructions above can be repeated at any time should you suspect the chart has faded or been damaged.

This chart was created by the University of Canberra, 11 Kirinari St, Bruce ACT 2617, Australia.

### Serif: Nimbus Roman

xxx − 0.10 xxx

xxx 0.00 xxx

xxx 0.10 xxx

xxx 0.20 xxx

xxx 0.30 xxx

xxx 0.40 xxx

xxx 0.50 xxx

xxx 0.60 xxx

xxx 0.70 xxx

xxx 0.80 xxx

xxx 0.90 xxx

| +0.90 logRAD | 6/48 | 0.125 | 3.2M | N24 |
|---|---|---|---|---|

Computer reading glasses are designed to reduce eye strain and keep things clear when doing screen work.

zones — vision — woven

| +0.80 logRAD | 6/38 | 0.16 | 2.5M | N18 |
|---|---|---|---|---|

Bifocal glasses have two zones: the top part for far away and the bottom part for reading. They allow clear vision at different distances.

screen — waxes — axons

| +0.70 logRAD | 6/30 | 0.20 | 2.0M | N14 |
|---|---|---|---|---|

Long periods of near work can cause headaches, tired eyes, and make it harder to keep things in focus. Regular breaks and good lighting can help reduce strain and improve comfort.

aroma — oceans — rescue

| +0.60 logRAD | 6/24 | 0.25 | 1.6M | N12 |
|---|---|---|---|---|

The brain and eyes work together to allow people to see. Although each eye sees a slightly different view, the brain combines the two views together to form one image.

nearer — sesame — wanes

UNIVERSITY OF CANBERRA

**UC/UWA Reading Chart**

Version 0.91.sans-serif (April 2026)

To be used at 40cm

Manufactured by the University of Canberra, Bruce ACT 2617, Australia

CC BY ND

Murphy TI, Chen J, Leung M. How accurate are our near reading charts? An assessment of 19 charts against ISO standards. medrxiv. 2026 Jan 30:2026-01.

THE UNIVERSITY OF WESTERN AUSTRALIA

**+0.50 logRAD | 6/19 | 0.32 | 1.3M | N10**

It is common for some people to see floaters, which can appear as dark or light spots or strings that move with the eye. If there are sudden increases in floaters, or flashing lights, this could indicate damage to the retina at the back of the eye which needs urgent attention.

sermon — common — assume

**+0.40 logRAD | 6/15 | 0.40 | 1.0M | N8**

For some people the front surface of the eye or the lens inside the eye has an uneven shape. This is called astigmatism and can cause people to see ghost images or shadows around objects up close and far away. Glasses or contact lenses can usually help make things clearer and reduce eye strain.

answer — secure — casino

**+0.30 logRAD | 6/12 | 0.50 | 0.80M | N6**

When the eye is too short it has to work harder to see things, especially up close. This is called long-sightedness or hyperopia, and can be tiring as the eye has to work harder than normal to see things clearly. Some people also get headaches after long periods of reading or screen time. Glasses can help to adjust the eye's focus, making close-up tasks like reading and sewing more comfortable.

ensure — sauna — waves

**+0.20 logRAD | 6/9.5 | 0.63 | 0.63M | N5**

People who can see clearly up close but not far away have short-sightedness. This is also known as myopia and is usually caused by the eyeball being too long. It normally develops in childhood and can worsen into early adulthood. The risk of eye problems increases as the eye grows, so it is important to manage myopia early. This can involve special contact lenses, glasses, or eyedrops.

arena — seasons — owner

**+0.10 logRAD | 6/7.5 | 0.80 | 0.50M | N4**

Inside the eye, there is a lens that helps focus light so that a person can see. A cataract is a normal change where the lens slowly becomes cloudy with age. Cataracts can cause vision to appear hazy and colours to appear less vibrant. In the early stages it can be managed with better lighting, but will need surgery once vision and quality of life are significant affected.

cannon — worms — arrow

**0.00 logRAD | 6/6.0 | 1.0 | 0.40M | N3**

As we get older, it becomes harder for the lens inside the eye to change shape, which makes reading small print more difficult. This is a normal change known as presbyopia and is not a sign of disease. Many people first notice it when holding a book or phone a little farther away than usual. Reading glasses or multifocal lenses can make near tasks clearer and more comfortable, with most people continuing their daily activities without any significant issues.

source — cursor — ransom

**−0.10 logRAD | 6/4.8 | 1.3 | 0.32M | N2**

The inside of the eye is lined with a layer of cells called the retina. These cells respond to light and enable us to see. As we get older these cells become less efficient, making it harder to adapt when moving from bright sunlight into a dark room. This is a natural change but can be quite frustrating when it affects everyday activities. Good lighting can make it easier for the eyes to adjust.

camera — reason — noise

UNIVERSITY OF CANBERRA

Conforms to ISO 7921:2024 when validated according to the accompanying documentation.

Validated by: ______________________

Signed: ______________________ Date: __________

THE UNIVERSITY OF WESTERN AUSTRALIA

### UC/UWA Reading Chart

#### Version 0.91.sans-serif (April 2026)

This reading chart conforms to ISO 7921:2024 provided the following checks are performed. It is the user's responsibility to ensure this has been completed in full before use in a clinical or research setting. The manufacturers make no guarantee of the accuracy of this reading chart, or conformance to ISO 7921:2024, should this procedure not be followed. Once all checks listed below have been completed, document that the chart has been validated by dating and signing the bottom of the back page in the space provided.

##### Printing

To ensure all text is printed legibly and in the correct size, this chart must be printed at a mimimum 1200 dpi on white, A4 paper or cardboard. This chart should have a matte surface to avoid reflections which may impact contrast. No additional margins or padding can be added as this will scale the text. This chart may be printed in colour or grayscale. Check legibility of the smaller sizes with a magnifying device.

##### Text size

The PDF version of this chart contains text of the correct size, within the margin of error allowed by the standard. To ensure accuracy, a validation page is printed on the reverse of this page. The rulers in the margins should be accurate and can be checked with a ruler. Each line displays the logRAD size with three 'x' characters on either side. The height of these characters should be within 5% of the corresponding x-height in the table below. Measurements must be done using a device with a measurement error of 0.01mm or less, such as callipers.

##### Contrast

Text must have a contrast of at least 85%. The PDF version of this chart has a 100% contrast but this may be reduced by the printing process. Contrast is calculated by measuring the luminance of the background surrounding the text ($L_b$) and luminance of the text itself ($L_t$). Background luminance must be between 80 and 200 cd/m$^2$ at a distance of 40cm (80-120 cd/m$^2$ recommended). The chart contrast is calculated as $C = \frac{L_b - L_t}{L_b}$. If $C \geq 0.85$ then the chart has sufficient contrast.

| **Reading acuity angle** min of arc | **x-height** mm | **Near reading acuity grades at 40cm** | | | | | |
|---|---|---|---|---|---|---|---|
| | | **logRAD**[a] | **Decimal reading acuity**[a,b] | **M size**[a] | **N size**[c] | **Reduced Snellen fraction** | |
| 20.0 | 11.61 | +1.30 | 0.05 | 8.00 | 60 | 20/400 | 6/120 |
| 15.8 | 9.22 | +1.20 | 0.063 (0.06) | 6.30 | 48 | 20/320 | 6/95 |
| 12.6 | 7.33 | +1.10 | 0.08 | 5.00 | 36 | 20/250 | 6/75 |
| 10.0 | 5.82 | +1.00 | 0.10 | 4.00 | 30 | 20/200 | 6/60 |
| 7.94 | 4.62 | +0.90 | 0.125 | 3.20 | 24 | 20/160 | 6/48 |
| 6.31 | 3.67 | +0.80 | 0.16 | 2.50 | 18 | 20/126 | 6/38 |
| 5.01 | 2.92 | +0.70 | 0.20 | 2.00 | 14 | 20/100 | 6/30 |
| 3.98 | 2.32 | +0.60 | 0.25 | 1.60 | 12 | 20/80 | 6/24 |
| 3.16 | 1.84 | +0.50 | 0.32 (0.3) | 1.30 | 10 | 20/63 | 6/19 |
| 2.51 | 1.46 | +0.40 | 0.40 | 1.00 | 8 | 20/50 | 6/15 |
| 2.00 | 1.16 | +0.30 | 0.50 | 0.80 | 6 | 20/40 | 6/12 |
| 1.58 | 0.922 | +0.20 | 0.63 (0.6) | 0.63 | 5 | 20/32 | 6/9.5 |
| 1.26 | 0.733 | +0.10 | 0.80 | 0.50 | 4 | 20/25 | 6/7.5 |
| 1.00 | 0.582 | 0.00 | 1.00 | 0.40 | 3 | 20/20 | 6/6.0 |
| 0.79 | 0.462 | −0.10 | 1.25 | 0.32 | 2 | 20/16 | 6/4.8 |
| 0.63 | 0.367 | −0.20 | 1.60 | 0.25 | 1.8 | 20/12.5 | 6/3.8 |
| 0.50 | 0.292 | −0.30 | 2.00 | 0.20 | 1.5 | 20/10 | 6/3.0 |

[a] Values that end in zero may be truncated to delete the last zero only for the purpose of identifying the near reading acuity grade.
[b] Values in parentheses shall be used only for the purpose of identifying the near reading acuity grade.
[c] Values are approximations for Helvetica and rounded to the nearest common size for the purpose of identifying the near reading acuity grade.

##### Use and care instructions

This chart is to be used at 40cm under ambient or task lighting. Light sources not intended to illuminate the chart should not create specular reflections, and light sources visible to the subject may not exceed the chart background luminance. To ensure the print quality of this reading chart, store away from direct sunlight. The validation instructions above can be repeated at any time should you suspect the chart has faded or been damaged.

This chart was created by the University of Canberra, 11 Kirinari St, Bruce ACT 2617, Australia.

### Sans Serif: Nimbus Sans

xxx − 0.10 xxx

xxx 0.00 xxx

xxx 0.10 xxx

xxx 0.20 xxx

xxx 0.30 xxx

xxx 0.40 xxx

xxx 0.50 xxx

xxx 0.60 xxx

xxx 0.70 xxx

xxx 0.80 xxx

xxx 0.90 xxx
